# CAIDE score, brain structure, and cognitive functions in middle-to-older aged adults: A KoGES population-based study

**DOI:** 10.64898/2026.05.19.26353376

**Authors:** Gisung Shin, Ali Tanweer Siddiquee, Seung-ku Lee, June Christoph Kang, Hyunsan Cho, Jisun Choi, Youngjin Kim, Bongjo Kim, Nanhee Kim, Chol Shin

**Affiliations:** Institute of Human Genomic Study, College of Medicine, Korea University Ansan Hospital, Republic of Korea; Institute of Brain Engineering, Korea University College of Informatics, Seoul, Republic of Korea; Division of Genome Science, Department of Precision Medicine, National Institute of Health, Chungcheongbuk-do, 28159, Republic of Korea; Division of Endocrinology and Metabolism, Department of Internal Medicine, Korea University Ansan Hospital, 123, Jeokgeum-ro, Danwon-gu, Ansan-si, Gyeonggi-do 15355, Republic of Korea

**Author notes:** Corresponding author: Nanhee Kim, MD, PhD, Institute of Human Genomic Study, College of Medicine, Korea University Ansan Hospital, #123, Jeokgeum-ro, Danwon-gu, Ansan, 15355, Republic of Korea, Chol Shin, MD, PhD, Institute of Human Genomic Study, College of Medicine, Korea University Ansan Hospital, #123, Jeokgeum-ro, Danwon-gu, Ansan, 15355, Republic of Korea.

## Abstract

**Background:** Although CAIDE (Cardiovascular Risk Factors, Aging, and Dementia) score estimates 20-year dementia risk, prior studies have largely focused on global or composite measures of cognitive function. Among the studies, only a few have investigated involving cognitive functions and structural neuroimaging markers, and the available structural neuroimaging evidence has largely been derived from subsamples or highly selected small cohorts rather than full population-based cohorts. We, therefore, not only investigated associations between CAIDE score and cognitive performance but also explored the mediating role of structural neuroimaging markers in middle-to-older aged population.

**Methods:** Of 2,864 participants who were available for structural magnetic resonance imaging (MRI) data at baseline, we excluded 230 participants who had neurological and cardiovascular disease at baseline. We also further excluded 209 participants without having exposure, covariates, and cognitive assessments data, and 2,425 participants were included for the final analysis. The CAIDE score was calculated based on midlife vascular risk factors. We categorized participants into low, moderate, and high-risk groups based on the tertile distribution of the total scores. For the original CAIDE score (without APOE4), the tertile-based cut-offs were <6, 6-to-7, and >7 points. In the sensitivity analysis incorporating APOE4 status, the cut-off points were recalculated to maintain tertile-based categorization, resulting in <7, 7-to-8, and >8 points for low, moderate, and high risk groups, respectively. The main outcomes were neuropsychological assessment battery included Story recall, Visual reproductions, Verbal fluency, Trail making, Digit symbol – coding, and Stroop tests.

**Findings:** Of 2,425 healthy participants (mean age of 58.5 [6.5]; men 1,189 [49.0]), higher CAIDE groups were associated with poorer cognitive performance across all cognitive domains. Compared with low-risk group, the high-risk group had significantly lower mean adjusted z-scores across all 12 cognitive assessments (all p <.001). The moderate-risk group also showed lower mean adjusted z-scores in all tests, except visual reproduction recognition and verbal fluency category tests.

**Interpretation:** This large-based population study showed the highest CAIDE risk group was independently associated with lower cognitive performance across all domains compared to the lowest risk group, suggesting the potential importance of managing these features for preserving neurological health in middle and older aged adults.

**Research in Context:** *Evidence before this study:* Recent studies reported the associations between CAIDE (Cardiovascular Risk Factors, Aging, and Dementia) score and cognitive functions in middle-to-older aged population. However, most studies investigated cognitive functions as composite measures or global score and the accompanying structural neuroimaging markers have not been well characterized. We therefore searched PubMed with “CAIDE risk score” [All fields] OR “dementia risk score” [All fields] OR “AD risk score” [All fields] AND “Neuroimaging” [All fields] OR “magnetic resonance imaging” [All fields] OR “brain volumetric” [All fields] AND “cognitions” [All fields] OR “cognitive functions” [All fields] AND “population-based” [All fields] between April 17^th^, 2006 to April, 17^th^, 2026 among CAIDE risk factors, brain volume, and cognitive functions in middle to older aged population. We identified only a few studies investigated on cognitive functions and structural neuroimaging markers, and the available structural neuroimaging evidence has largely been derived from subsamples or highly selected small cohorts rather than full population-based cohorts. We therefore examined the associations between CAIDE score and cognitive functions across all domains and further explored the potential role of structural brain measures in middle to older-aged East Asian population.

*Added value of this study:* Notably, this large middle-to-older aged (49 to 80 years) population study extends from prior evidence by examine the associations between CAIDE score and cognitive functions (logical memory, visual reproduction, verbal fluency, trail making test A, digit symbol – coding, and stroop tests) rather than relying solely on composite measures or global score of cognitive functions. Additionally, we further explore structural brain volumes whether underlying these associations. We further found the structural neuroimaging measures alone did not fully account for the associations between CAIDE score and cognitive functions, suggesting a more complex relationship.

*Implications of all the available evidence:* This large population based study highlights the importance of CAIDE risk profiles managements before onset of the cognitive impairment. Notably, our results are consistent with prior studies reporting associations between the CAIDE score and both cognitive function and structural neuroimaging measures. Importantly, we extended these findings by demonstrating that structural brain measures may partially account for the association between CAIDE score and cognitive function in this asymptomatic middle-aged population. Overall, our results suggest that the association between the CAIDE score and cognitive performance may not be fully explained by macroscopic structural brain measures only. Thus, future studies are warranted to examine additional neurobiological mechanisms beyond structural brain measures that may contribute to the associations between CAIDE risk profile and cognitive function.

## Introduction

According to world health organization (WHO), more than 55 million people worldwide are living with dementia, with nearly 10 million new cases each year, making it a leading cause of death among older adults.^1^ By 2050, the global burden of dementia is projected to nearly triple.^2^ While dementia had been publicly well recognized as health concerns, cognitive impairments represent one of the most public health challenges in aging populations.

The cardiovascular risk factors, aging, and dementia (CAIDE) score, developed from a Finnish population-based cohort, incorporates common risk factors including age, sex, education, body mass index (BMI), systolic blood pressure (SBP), total cholesterol (TC), and physical activity to estimate long-term dementia risk.^3^ Apolipoprotein E (APOE) is also well known risk factor for dementia risk, especially in Alzheimer’s disease.^4^ Emerging evidence suggests that vascular and genetic risk factors may contribute to subclinical cognitive deficits and structural brain alterations even before progression to symptomatic dementia.^5^

Prior studies (including the Finnish Geriatric Intervention Study to Prevent Cognitive Impairment and Disability [FINGER] and AlzEpi Cohort Observational Library [ACOL] studies) have demonstrated associations between CAIDE score and cognitive performance as well as structural brain measures. However, neuroimaging evidence has largely been derived from subsamples^6^ or highly selected small cohorts^7^ rather than full cohorts. Accordingly, despite the established contribution of vascular and genetic risk factors to dementia development, their associations with cognitive and structural brain measures in asymptomatic middle-to older aged populations have not been fully characterized. To address these gaps, this study examines how the accumulated vascular burden, captured by the baseline CAIDE score, is reflected in brain structure and cognitive performance in asymptomatic middle-to-older aged adults.

## Methods

### Study Participants

Since 2001, Korean Genome and Epidemiology Study (KoGES) was initiated in Ansan, South Korea.^8^ In 2011 – 2014, KOGES Ansan aging study was launched with additional core examination of neuropsychological assessment battery and magnetic resonance imaging (MRI) at Exam 6 (2011-2012) and Exam 7 (2013-2014).^9^ Among 2,864 study participants who underwent MRI (aged from 49 to 80 years old) at baseline, 2,634 participants were free of neurological (dementia n=2; cerebrovascular disease n=90) and cardiovascular disease (n=138). We further excluded 21 participants who had missing exposure (education n=4 and systolic blood pressure n=1) and covariates (beck depression inventory [BDI] n=16) information and incomplete cognitive assessments (n=188). We finally analyzed 2,425 participants who completed both structural neuroimaging markers and neuropsychological assessments at baseline (2011 – 2014) in Figure 1.

**Figure 1.**
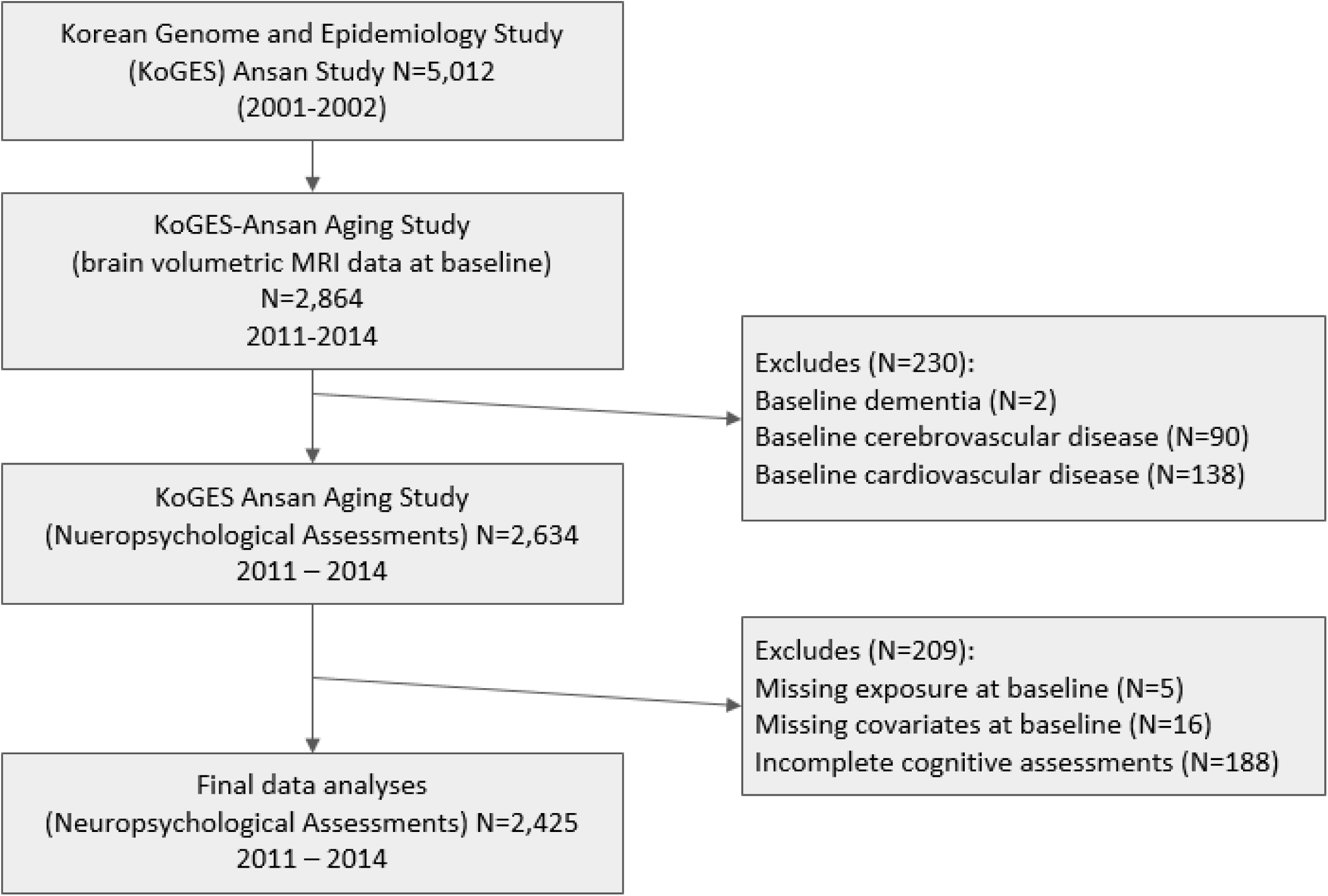
KoGES Ansan cohort study participants flow chart. The flowchart shows the stepwise exclusion of participants based on history of neurological disease, no information on variables (including age, sex, education level, body mass index, physical activity, systolic blood pressure, and total cholesterol) that included in CAIDE scores and confounding variables (including smoking and drinking status, a history of diabetes, age-related white matter changes). We further excluded incomplete cognitive assessments at baseline. We finally analyzed with 2,425 participants at baseline (2011-2014).

This study was approved by the Institutional Review Board of Korea University Ansan Hospital (IRB NO 2006AS0045) and written informed consent was obtained from all study participants. We followed the Strengthening the Reporting of Observational Studies in Epidemiology (STROBE) guidelines for reporting this observational study.

### CAIDE score assessments

CAIDE score was assessed with age, sex, education, BMI, SBP, TC, and physical activity. BMI was calculated as weight divided by height squared (kg/m^2^). Physical activity (PA) was assessed using a self-reported questionnaire regarding regular leisure-time physical activity. Information regarding the frequency (per week) and duration (min) of leisure time PA during a typical week was collected.^10^ Physical activity was defined as at least twice per week and at least 30 minutes per session of exercise. Blood pressure was measured in both arms using an appropriately sized cuff and a mercury sphygmomanometer (Baumanometer-Standby; W.A. Baum Co. Inc., New York, USA) after participants had rested in a seated position for at least 5 minutes. Both systolic and diastolic blood pressure were defined as the average of the left and right arm readings.^11^ The mean systolic blood pressure was used to derive the blood pressure component of the CAIDE score. All blood samples were obtained after at least 8 hours overnight fast and were immediately stored at −80°C for subsequent assays. 7600 Chemistry Analyzer (Hitachi; Tokyo, Japan) enable to enzymatically measure plasma concentrations including TC. Additionally, Apolipoportein E (APOE) genotype was determined as part of a large-scale analysis using the Korea Biobank Array (KoreanChip), a microarray platform optimized for the Korean population containing over 833,000 genetic markers (rs429358 and rs7412).^12^

APOE4 carrier status defined as at least one ε4 carrier (ε2/ε4, ε3/ε4, or ε4/ε4) then considered as an ε4 carrier, else non-carrier. Although the original CAIDE score is a validated tool designed to predict dementia risk based on midlife cardiovascular profiles, we used a modified version of the CAIDE score to reflect risk factor distributions and clinical characteristics in our study populations.

In this study, the CAIDE score was calculated according to original CAIDE score^3^, with a total score ranging from 0 to 15 points (pts). The score includes age, sex, education, BMI, SBP, TC, and physical activity as shown in eTable 1a. Age was scored as 0 points for <47 years, 3 pts for 47-53 years, and 4 pts for >53 years. Although the 0-point category was not represented in our study population, the original score was retained to preserve the relative contribution of age within the total score. Education level was categorized into three groups according to years of education: 0-6 years, 7-9 years, and 10 or more years corresponding to elementary school or less, middle school, and high school or higher education in the Korean educational system. BMI was categorized using a cutoff 25 kg/m^2^ according to Korean Society for the study of Obesity (KSSO) criteria^13^ for Korean population. The total CAIDE score was further categorized into three groups (low [<6 pts], moderate [6-7 pts], and high risk [>7 pts]) based on tertile of the score distribution in our study population to examine associations across increasing levels of risk. In sensitivity analyses including APOE4 carrier status, the scoring criterial were expanded and the tertile-based cut-off points were recalculated accordingly to maintain the relative risk distribution, as shown in eTable 1b. Although the original CAIDE dementia risk score ranges from 0 to 18 points with APOE4 status, observed scores in our study populations were ranged from 0 to 16.

### Neuropsychological Assessment Battery

KoGES neuropsychological tests^14^ was adapted from the protocols used in the original and offspring cohorts of the Framingham Heart Study (FHS). Standardized and validated Story Recall (SR) tests were used to evaluated verbal memory with analogous to the Wechsler Memory Scale – Third Edition (WMS III).^15^ Verbal Fluency (VF) assessments consists two assessments: phonemic (VF1) and categorical fluency (VF2), which derived from the Controlled Oral Word Association Test.^16^ Visual memory evaluated using Visual Reproduction (VR) test from the Wechsler Memory Scale – Revised. Digit Symbol – Coding (DS) used for visual processing and sustained attention evaluation from the Wechsler Adult Intelligence Scale – Fourth edition guidelines. Simple attention assessed with Trail Making Test (TMT-A; number of sequencing) and Stroop Test Word Reading (Stroop 1). Executive function assessed with Stroop Test Color Reading (Stroop 2).^15–18^ All cognitive test scores were converted into z-scores (mean=0, standard deviation [SD]=1) based on overall study population. This allowed direct comparison of the effect size across cognitive domains. This neuropsychological assessment battery emphasizes higher z-scores as better performance, except TMT-A.

### Magnetic Resonance Imaging (MRI)

All 3D T1 weighted MRI data scans were performed with GE Signa 1.5 T MR scanner (GE Medical Systems, Waukesha, WI, USA) with an 8-channel head coil from March 4, 2011 to January 26, 2013. The 3D T1-weighted images were acquired with the following parameters (TR=7.7ms, TE=3.4ms, flip angle=12°, slice thickness=1.6mm). Structural MRI data were processed using the BRAINS Auto-Workup pipeline^19^, which incorporates SyN-based nonlinear registration for accurate tissue segmentation and atlas-based regional volume estimation. All volumetric measurements, including gray matter (GM), white matter (WM), and cerebrospinal fluid (CSF), were extracted in each individual’s original anatomical space, without any distortion introduced by warping to an arbitrary template. Brain volumes (GM, WM, and CSF) were divided by intracranial volume (ICV) to account for individual differences in head size and then standardized to z-scores.

### Covariates

KoGES questionnaires included demographic, lifestyle, and history of disease status were biennially collected data with a standardized manual. Both self-reported alcohol and smoking status were categorized as never, past, and current. The BDI score is used to assess depressive severity. Each item is rated on a four point Likert scale (0 – 3), with total scores ranging from 0 to 63.^20^ Type 2 diabetes (DM) defined as fasting blood ≥ 126 mg/dL or usage of insulin or oral hypoglycemic agents’ and categorized into binary variables based on medical history and current medication use.^21^ Age-related white matter changes (ARWMC) were assessed based on T2-weighted FLAIR (Fluid-Attenuated Inversion Recovery) images using GE Signa 1.5 T MR scanner (GE Medical Systems, Waukesha, WI, USA). Participants were classified as having ARWMC (score ≥1) or none to reduce model complexity. We thereby included this variable as a covariate to account for potential confounder.^22^

### Statistical Analysis

Continuous variables were presented as mean (standard deviations [SD]) for normally distributed data and compared using one-way analysis of variance (ANOVA), or median (interquartile range [IQR]) for non-normally distributed data and compared using the Kruskal-Wallis test.

Categorical variables were presented as counts (percentages) and compared using chi-square test. To examine the association between CAIDE groups and cognitive function, we used multivariable linear regression models using PROC GLM procedures in SAS 9.4 (SAS Institute, Cary, NC, USA) to estimate adjusted mean differences in z-score, with the low-risk group (<6 pts) as the reference. To explore the potential indirect association between CAIDE score and cognitive functions through brain volumes (GM, WM, and CSF), we conducted separate mediation models within a path analysis framework. Indirect effects were estimated using a bootstrapping procedures with 5,000 resampling methods.^23^ Mediation analysis were performed using pingouin (version 0.5.5) package and covariate adjustments were conducted using statsmodels (version 0.14.4) procedures in python 3.9 (Python Software Foundation, Delaware, USA). Standardized brain volumes and cognitive scores were used in these cross-sectional models. To evaluate the robustness of our primary findings, we conducted sensitivity analyses. First, we incorporated APOE4 carrier status into the CAIDE score as genetic risk component (eTable 1a and 1b), given that APOE4 is the well-known established genetic risk factor for Alzheimer’s disease and has been incorporated into extended versions of the CAIDE score.^3,24^ Second, we repeat the primary analysis by combining the low-to-moderate risk groups to compare with the high-risk group. A two tailed p-value <0.05 was considered statistically significant. All statistical analyses were performed with SAS 9.4 software (SAS Institute, Cary, NC, USA) and python 3.9 (Python Software Foundation, Delaware, USA).

## Results

### Baseline characteristics

Table 1 shows the baseline characteristics of study participants (n=2,425) across CAIDE groups. The highest CAIDE groups were generally older, more proportionately in men, less educated, more obese, less levels of exercise, and had higher levels of total cholesterol and hypertensive (higher SBP/DBP, TG, and FBG but lower HDL). The highest CAIDE groups were also more likely to be diabetic and had proportionately higher presence of ARWMC (p <.001). Additionally, significant differences in relative gray matter, white matter, CSF volumes were observed among CAIDE groups, with the highest CAIDE score showed lower relative gray matter and white matter volumes and higher relative CSF volume in Table 1.

**Table 1.**
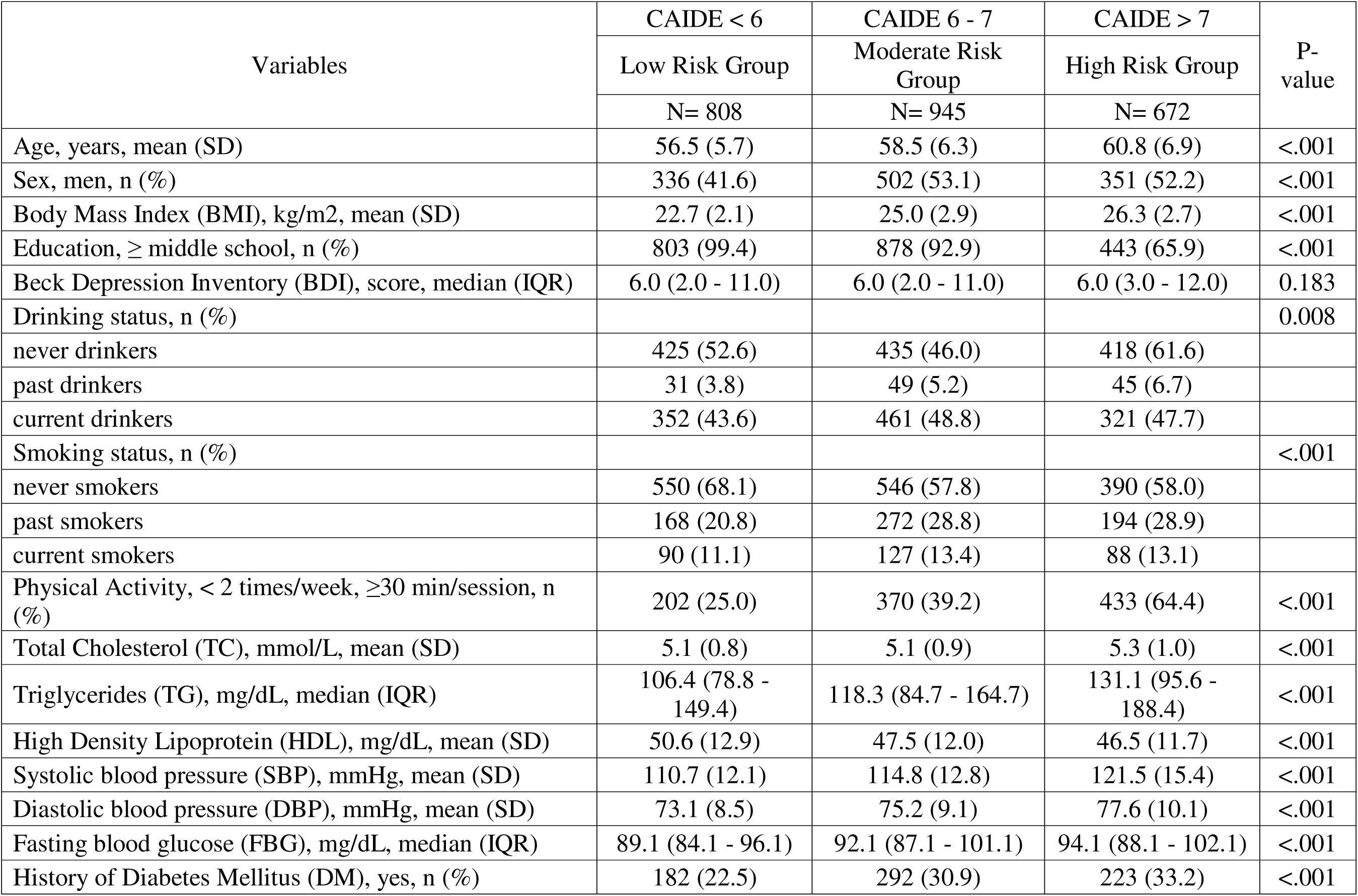
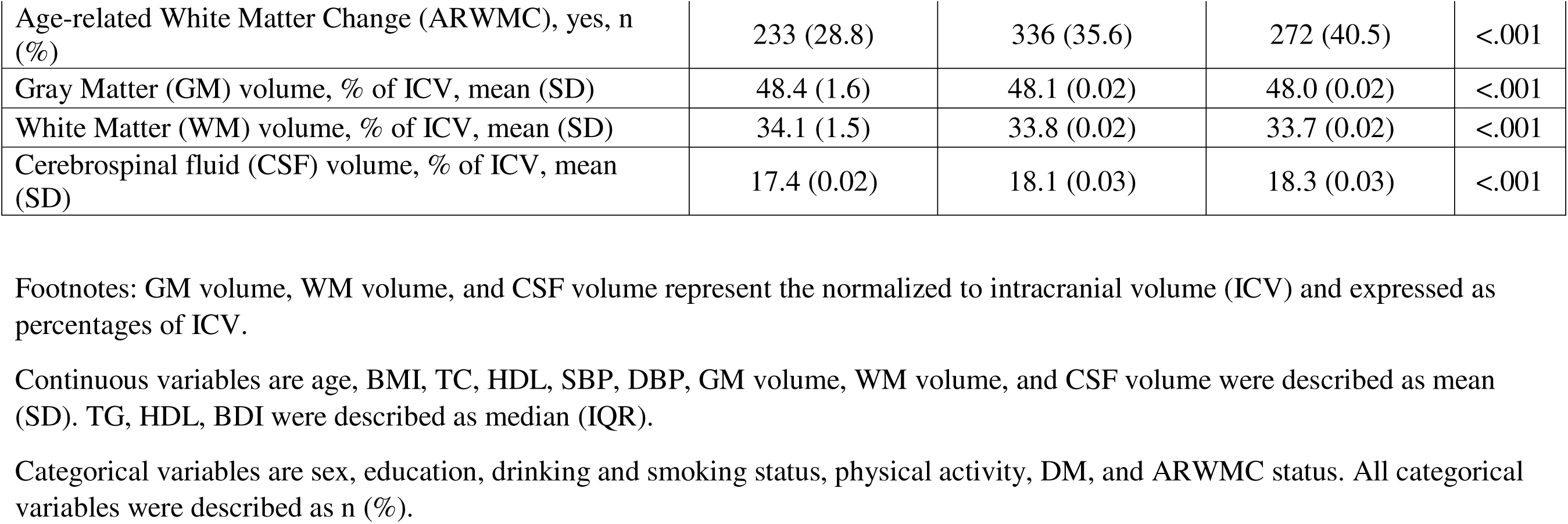
Baseline characteristics of the study participants (N=2,425) by CAIDE groups (low, moderate, and high risk groups).

### Association between CAIDE groups and cognitive performance

The highest CAIDE group was significantly different compared to the lowest risk group in all of the 12 neuropsychological tests at baseline (Figure 2, Table 2). Compared to the low-risk group, high risk group had lower cognitive performance in logical memory immediate (adjusted β estimates −0.445, 95% confidence intervals (CI): −0.566 to −0.323, p<.001) and delayed recall (adjusted β estimates −0.473, 95% CI: −0.594 to −0.352, p<.001) and recognition (adjusted β estimates −0.430, 95% CI: −0.552 to −0.309, p<.001), visual reproduction immediate (adjusted β estimates −0.581, 95% CI: −0.699 to −0.462, p<.001) and delayed recall (adjusted β estimates −0.544, 95% CI: −0.663 to −0.425, p<.001) and recognition (adjusted β estimates −0.489, 95% CI: −0.609 to −0.369, p<.001), verbal fluency phonemic (adjusted β estimates −0.567, 95% CI: −0.686 to −0.448, p<.001) and category (adjusted β estimates −0.374, 95% CI: −0.495 to −0.253, p<.001), trail making test A (adjusted β estimates 0.588, 95% CI: 0.470 to 0.705, p<.001), digit symbol coding test (adjusted β estimates −0.916, 95% CI: −1.028 to −0.803, p<.001), and stroop word reading (adjusted β estimates −0.719, 95% CI: −0.837 to −0.603, p<.001) and color reading (adjusted β estimates −0.526, 95% CI: −0.659 to −0.422, p<.001). Moderate risk group had lower cognitive performance in logical memory immediate recall (adjusted β estimates −0.170, 95% CI: −0.281 to −0.059, p=.001), delayed recall (adjusted β estimates −0.177, 95% CI: −0.287 to −0.066, p<.001), and recognition (adjusted β estimates −0.182, 95% CI: −0.293 to −0.071, p<.001), visual reproduction immediate recall (adjusted β estimates −0.186, 95% CI: −0.294 to −0.077, p<.001), delayed recall (adjusted β estimates –0.179, 95% CI: −0.288 to –0.070, p<.001), verbal fluency phonemic (adjusted β estimates −0.208, 95% CI: −0.317 to −0.099, p<.001), trail making test –A (adjusted β estimates 0.200, 95% CI: 0.093 to 0.307, p<.001), digit symbol – coding (adjusted β estimates −0.326, 95% CI: −0.428 to −0.223, p<.001), and stroop word reading (adjusted β estimates −0.248, 95% CI: −0.355 to −0.141, p <.001) and color reading (adjusted β estimates −0.206, 95% CI: −0.314 to –0.097, p<.001) compared to low risks group by CAIDE score. Except visual reproduction recognition and verbal fluency category tests in moderate risks were not significantly different from low-risk groups.

**Figure 2.**
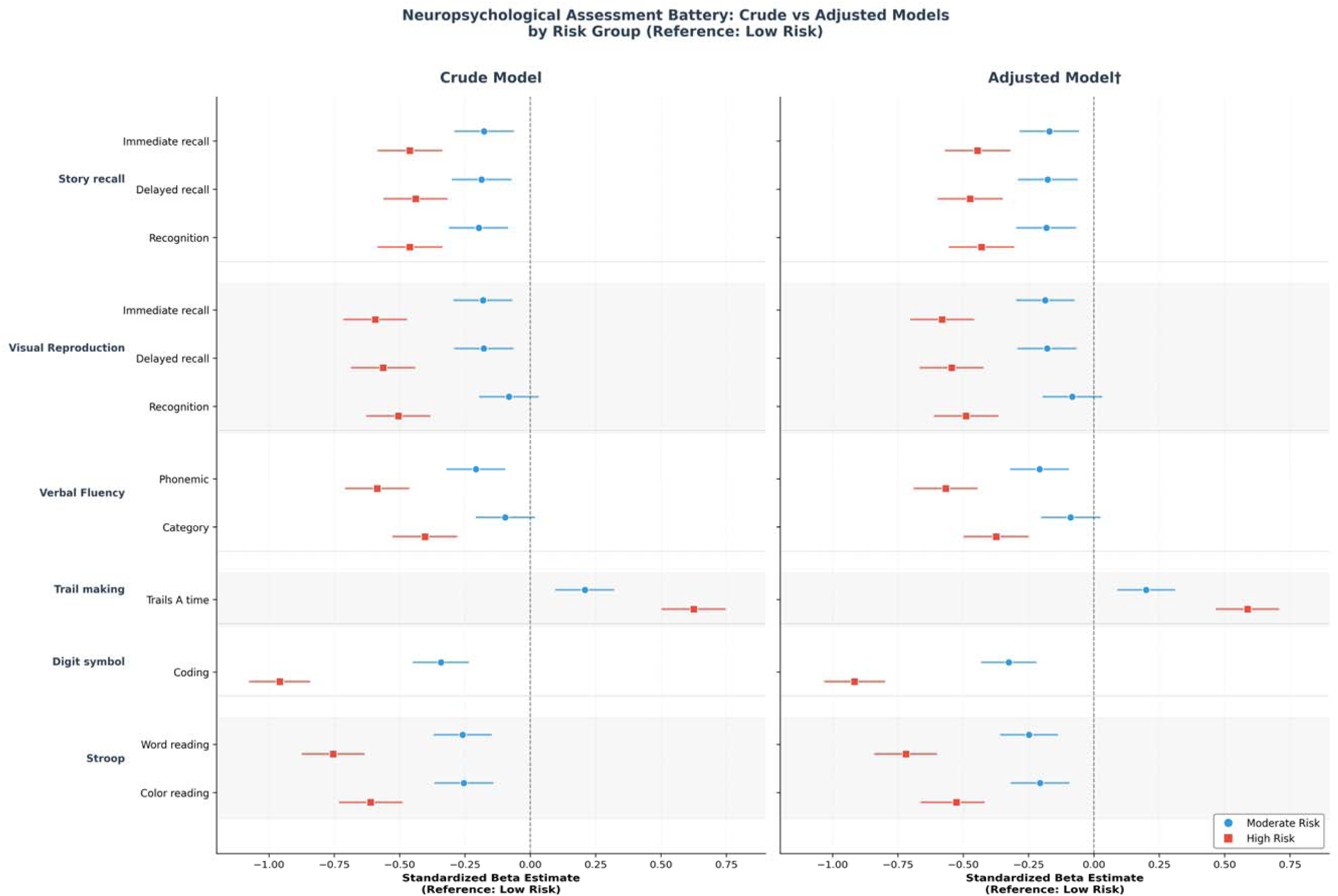
Multivariable adjusted β estimates and 95% confidence intervals of cognitive performance associated with CAIDE groups at baseline in the forest plot (N = 2,425). The low-risk group was used as references. Negative β estimates emphasize lower cognitive performance compared with the low-risk group except TMT-A. Circles represent moderate risk group (6-7) and squares represent high risk group (>7) after multivariable analyses. We adjusted for smoking and drinking status, BDI, DM, ARWMC.

**Table 2.**
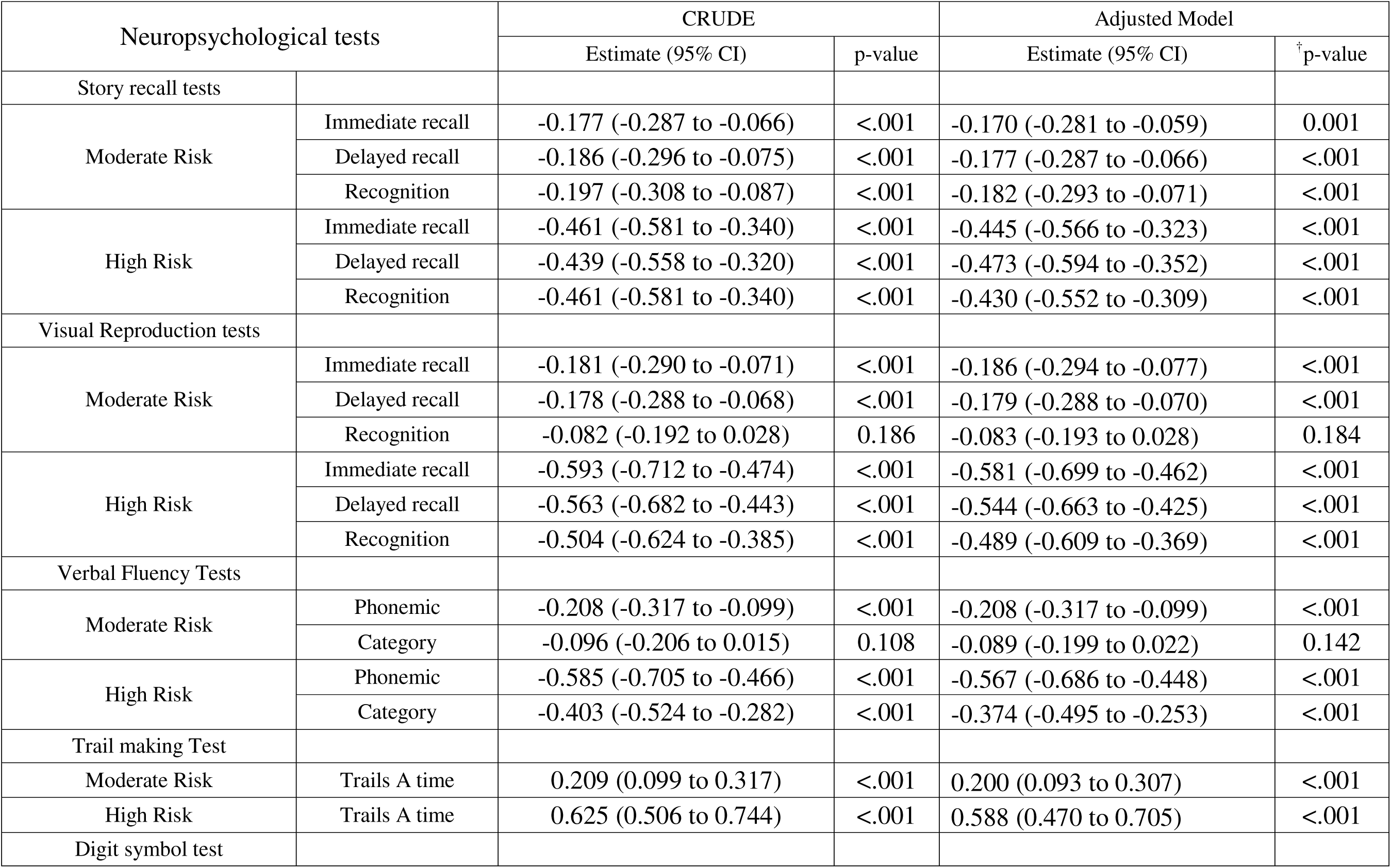
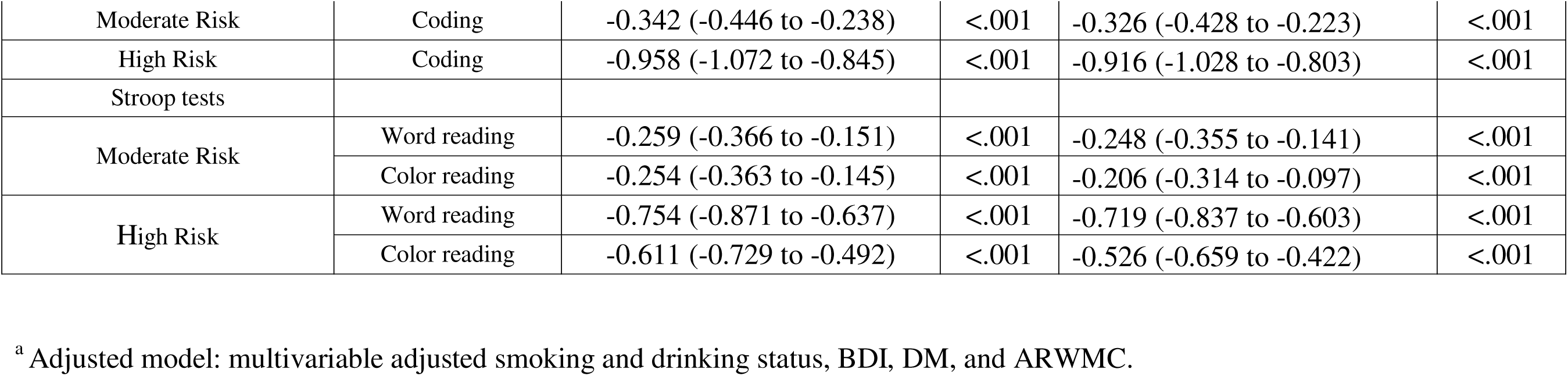
Multivariable adjusted^a^ mean difference (95% Cl) of cognitive function test scores using general linear model with analysis of covariance (ANCOVA) among CAIDE groups (N = 2,425).

In secondary analyses, GM, WM, and CSF volumes all partially mediated the association between higher CAIDE scores and lower cognitive performance. Higher CAIDE scores were associated with lower WM and GM volumes as well as higher CSF volume, which linked to lower cognitive scores across multiple domains. Across the multiple cognitive domains examined, the proportion of the total effect mediated (PM) ranged from approximately 1 to 3% for GM volume, 1% to 5% for WM volume, and 1% to 7% for CSF volume even after adjusted for smoking and drinking status, BDI score, DM and ARWMC status (Figure 3).

**Figure 3.**
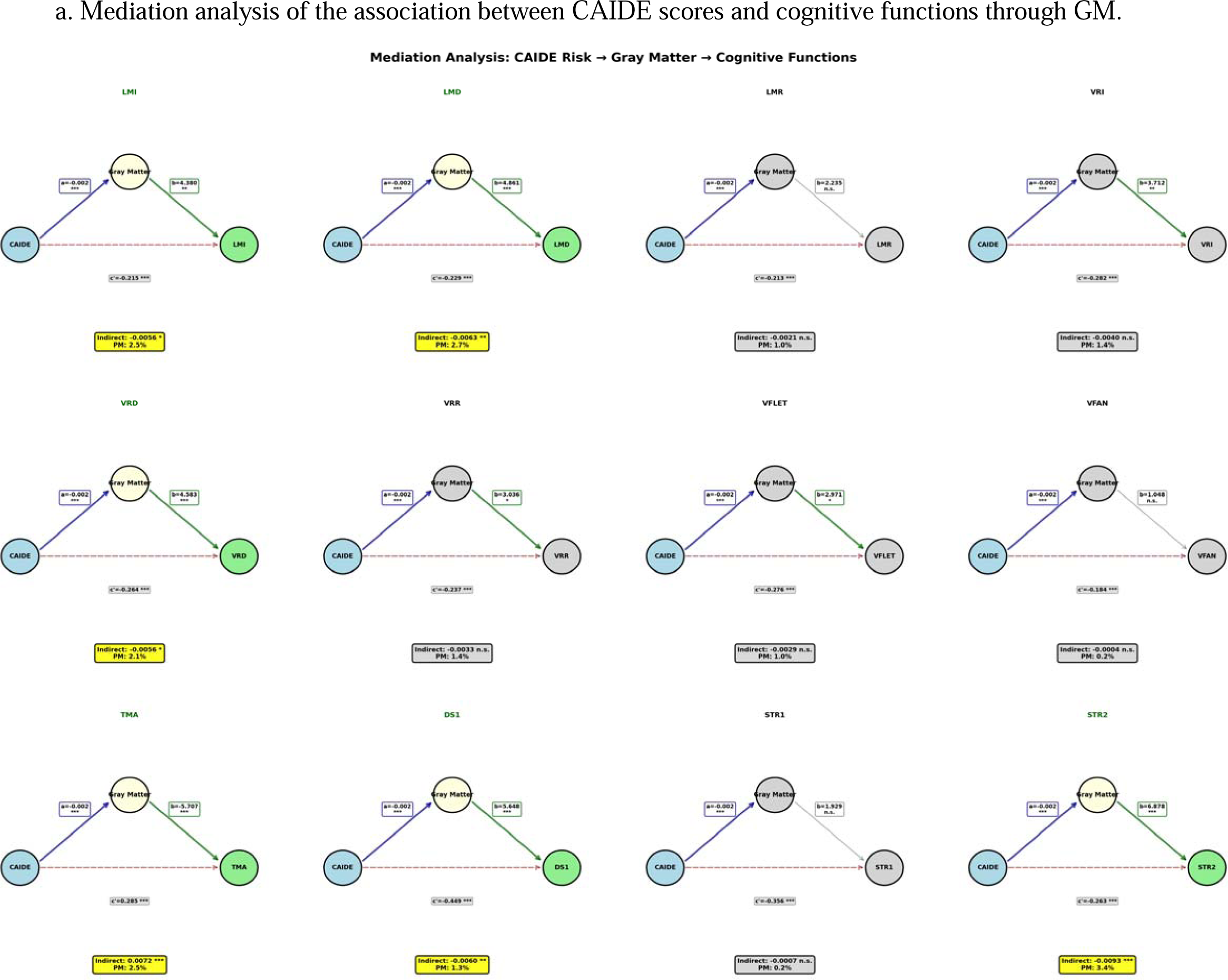
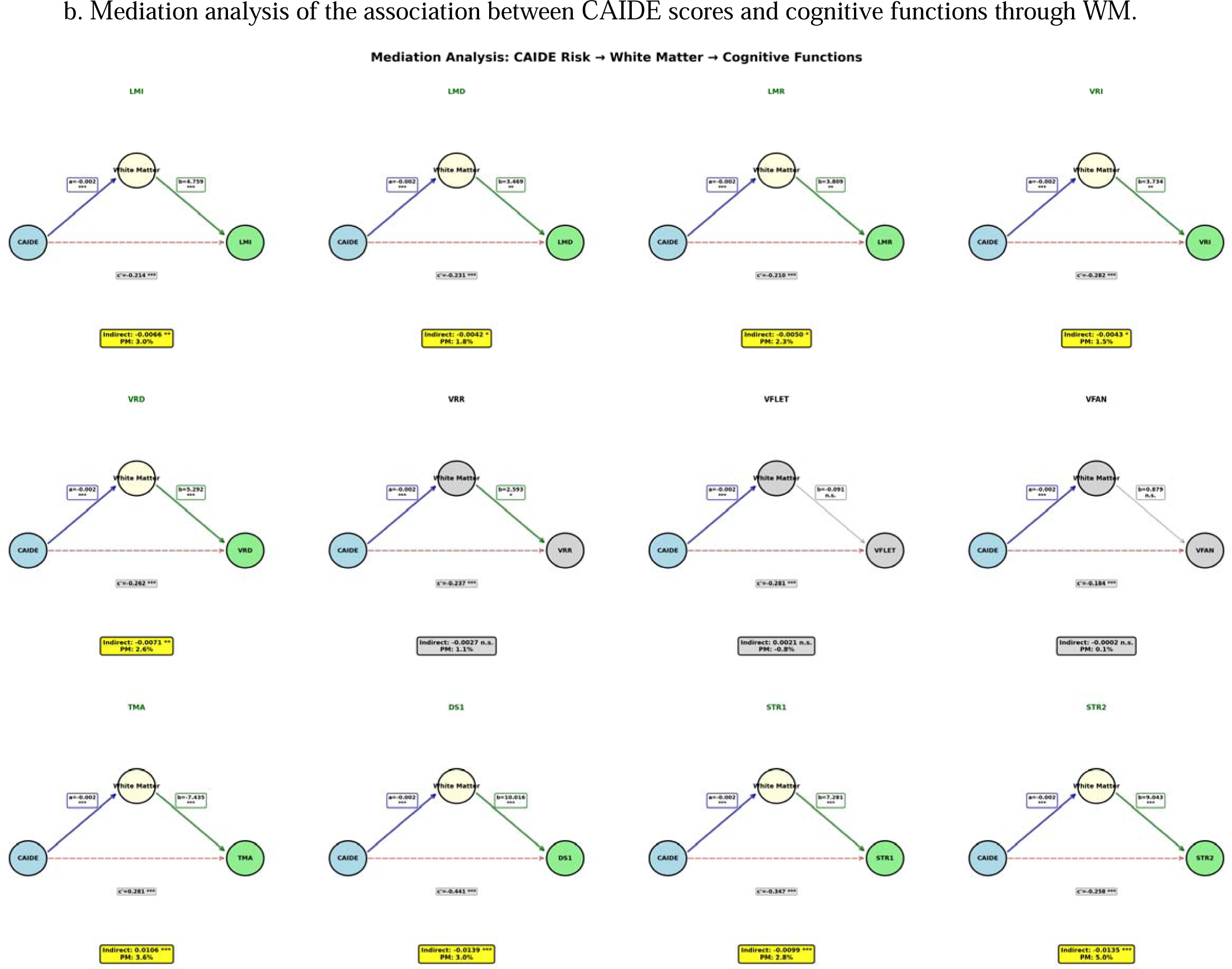
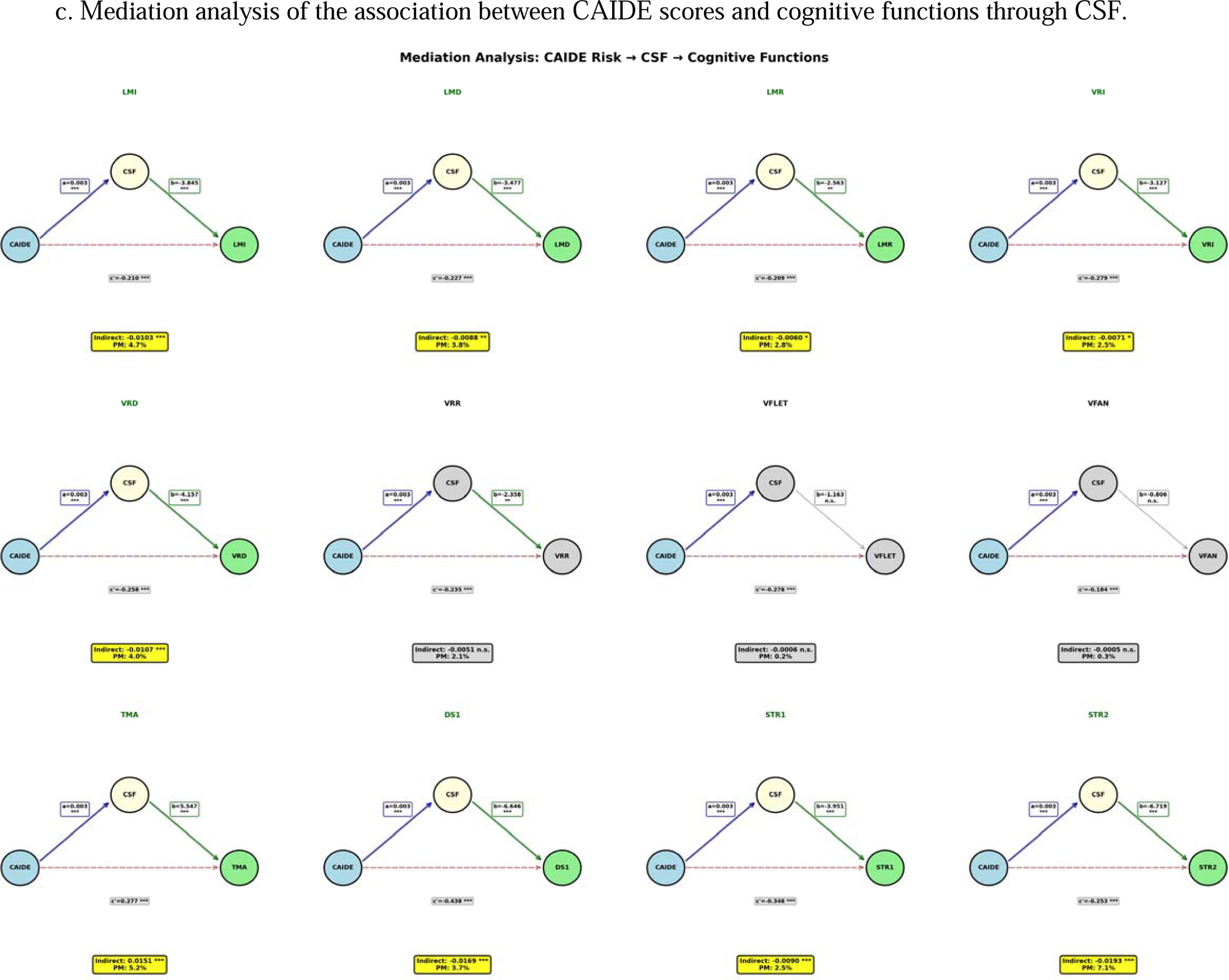
Cross-sectional mediation models examining whether brain volumes (GM, WM, and CSF) statistically incorporate associations between CAIDE score and cognitive performance. Path coefficients and significance levels are presented for each pathway: (A) GM volume, (B) WM volume, and (C) CSF volume. Significant indirect effects (p<0.05) are highlighted in yellow boxes. Analyses were additionally adjusted for smoking, alcohol status, BDI, diabetes status, and ARWMC as potential confounder factors associated with cognitive functions. Significance levels represents as followed: *p<.05 (*), p<.01 (**), and p<.001 (***)*.

### Sensitivity Analyses

First, although some associations were attenuated in sensitivity analyses incorporating baseline APOE4 carrier status (n=2,063) into the original CAIDE model, the results were generally similar (eTable 2). Compared with low-risk group, logical memory immediate (adjusted β estimates −0.405, 95% confidence intervals (CI): −0.530 to −0.280, p<.001), delayed recall (adjusted β estimates −0.441, 95% CI: −0.565 to −0.316, p<.001) and recognition (adjusted β estimates −0.382, 95% CI: −0.506 to −0.257, p<.001), visual reproduction immediate (adjusted β estimates −0.501, 95% CI: −0.623 to −0.379, p<.001) and delayed recall (adjusted β estimates −0.512, 95% CI: −0.634 to −0.391, p<.001) and recognition (adjusted β estimates −0.438, 95% CI: −0.562 to −0.314, p<.001), verbal fluency phonemic (adjusted β estimates −0.495, 95% CI: −0.617 to −0.373, p<.001) and category (adjusted β estimates −0.325, 95% CI: −0.449 to −0.201, p<.001), trail making test A (adjusted β estimates 0.569, 95% CI: 0.450 to 0.689, p<.001), digit symbol coding test (adjusted β estimates −0.859, 95% CI: −0.973 to −0.745, p<.001), and stroop word reading (adjusted β estimates −0.663, 95% CI: −0.782 to −0.543, p<.001) and color reading (adjusted β estimates −0.542, 95% CI: −0.663 to −0.422, p<.001) were lower in high risk group. Compared with low risk group, the moderate risk group showed significantly lower cognitive performance in logical memory immediate recall (adjusted β estimates −0.141, 95% CI: −0.269 to −0.012, p=0.028), delayed recall (adjusted β estimates –0.128, 95% CI: −0.256 to 0.001, p=0.051), and recognition (adjusted β estimates −0.137, 95% CI: −0.265 to −0.009, p=0.033), trail making test A (adjusted β estimates 0.155, 95% CI: 0.032 to 0.278, p=0.009), digit symbol – coding (adjusted β estimates −0.227, 95% CI: −0.344 to −0.110, p<.001), and stroop word reading (adjusted β estimates −0.179, 95% CI: −0.302 to −0.056, p=0.002) and color reading (adjusted β estimates −0.187, 95% CI: −0.311 to –0.063, p =0.001). No significant differences were observed for overall visual reproduction and verbal fluency tests in moderate risk groups (eTable 2).

Second, in sensitivity analyses combining the low and moderate risk groups and comparing them with the high risk group, the results were comparable to those of the primary analyses (eTable 3). Compared with the combined low and moderate risk group, the high risk group showed significantly lower cognitive performance in logical memory immediate (adjusted β estimates −0.351, 95% confidence intervals (CI): −0.439 to −0.263, p<.001), delayed recall (adjusted β estimates −0.376, 95% CI: −0.464 to −0.288, p<.001) and recognition (adjusted β estimates −0.329, 95% CI: −0.418 to −0.242, p<.001), visual reproduction immediate (adjusted β estimates −0.479, 95% CI: −0.565 to −0.393, p<.001) and delayed recall (adjusted β estimates −0.445, 95% CI: −0.531 to −0.359, p<.001) and recognition (adjusted β estimates −0.444, 95% CI: −0.531 to −0.357, p<.001), verbal fluency phonemic (adjusted β estimates −0.452, 95% CI: −0.539 to −0.366, p<.001) and category (adjusted β estimates −0.325, 95% CI: −0.412 to −0.238, p<.001), trail making test A (adjusted β estimates 0.477, 95% CI: 0.392 to 0.562, p<.001), digit symbol coding test (adjusted β estimates −0.736, 95% CI: −0.818 to −0.654, p<.001), and stroop word reading (adjusted β estimates −0.583, 95% CI: −0.668 to −0.498, p<.001) and color reading (adjusted β estimates −0.428, 95% CI: −0.513 to −0.342, p<.001).

## Discussion

This large population based study demonstrated higher CAIDE risk groups were associated with poorer cognitive performance across multiple cognitive domains (including verbal and visual memory, language processing, processing speed, and executive functions) in middle and older aged adults. Additionally, higher CAIDE scores were also indirectly associated with lower WM volume, GM volume and higher CSF volume. This study extends on the existent limited evidence by demonstrating that higher CAIDE score is independently associated with lower domain-specific cognitive performance and further explored associated mediating roles of structural neuroimaging markers in a large, middle-to-older aged East Asian population.

Prior studies (including FINGER, Multi-Ethnic Study of Atherosclerosis [MESA], and Whitehall II study)^6,25,26^ have reported associations between higher CAIDE scores and poorer cognitive performance across several domains, including executive function, processing speed, and memory. However, the cognitive domains associated with higher CAIDE scores have varied across studies. Whitehall II study reported greater decline in global cognition (including reasoning and vocabulary) but not memory. In contrast, FINGER study reported associations across multiple domains (including executive function, processing speed, and memory) whereas MESA demonstrated associations primarily with global cognition (cognitive abilities screening instruments [CASI]), processing speed, and attention/working memory. These differences may partly reflect variations in population characteristics, study design, and cognitive assessment methods. Because cognitive performance was assessed at a single time point in our study, our findings are less likely to be influenced by practice effects that may affect repeated cognitive testing in longitudinal studies (Figure 2).^27^ Recent studies have further examined these relationships in more specific contexts. Aiello et al. (2025)^5^ reported associations between higher CAIDE scores and poorer cognitive performance in limited cognitive measures in younger population without neuroimaging data, while Farkas et al. (2025)^7^ observed associations with both cognitive performance and structural brain volumes but did not formally evaluate the extent to which neuroimaging markers accounted for these associations. In clinical populations, Enache et al (2016)^28^ also reported associations between higher CAIDE scores and biomarkers of neurodegeneration in memory clinic patients without dementia. However, these findings were derived from study participants from clinics which may not be directly generalizable to our asymptomatic population. In addition, neuroimaging data in large cohort study such as FINGER have often been limited to subsamples rather than the full study population. Our findings are broadly consistent with prior studies reporting associations between higher CAIDE scores and poorer cognitive performance across multiple cognitive domains (Figure 2, Table 2). Notably, our study extends the existing literature by providing a more detailed characterization of cognitive performance and by further exploring structural neuroimaging markers within a large middle to older aged population.

Extending these findings, our results suggest that the association between CAIDE score and poorer cognitive performances may not be fully explained by macroscopic brain structural differences only. Although overall structural brain volumes were associated with both CAIDE score and cognitive performance, they accounted for only a small proportion (approximately 1-7%) of the total effect (Figure 3a, 3b, and 3c). Notably, these structural pathways were served as partial mediators, the strength and consistency of these structural pathways differed by metric. WM and CSF volumes showed indirect associations with cognitive performance across a broader range of domains compared to GM volumes. This difference may reflect the heightened sensitivity of WM volume and increased CSF volume to early vascular-related neurodegeneration, whereas GM volume may less sensitive to early-stage of vascular risk factors compared to WM and CSF volume in this asymptomatic, middle to older aged population (eFigure 1). The accumulated vascular burden reflected by CAIDE score may exert a more pronounced structural impact on white matter and CSF metrics compared to GM volume in our multivariable-adjusted model, independent of visible small vessel disease burden. This structural specificity is further supported by the univariable correlation matrix of CAIDE components (eFigure 1) and further aligns with a prior longitudinal evidence showing that higher educational attainment does not alter the rate of structural brain aging.^29^ In our cohort, while higher age and SBP were associated with lower brain volumes at baseline, education showed virtually no direct correlation with macroscopic structural brain volumes. This contrasting patterns, where dynamic vascular risk factors correlated with lower overall structural brain volumes (GM, WM, and CSF) while education showed no such structural association despite its strong link to cognitive outcomes (eFigure 1), strongly suggest that education may operate primarily through a ‘passive reserve’ mechanism (an initial cognitive advantage) rather than directly modifying the underlying macroscopic neural structures. Accordingly, the lower cognitive performance among individuals with lower education, despite the absence of macroscopic brain atrophy, suggests that the CAIDE score may capture subclinical neurocognitive vulnerability in this middle-to-older aged asymptomatic population based study. The observed associations remained consistent across multiple sensitivity analyses (including the use of an extended CAIDE score incorporating APOE4 status and alternative risk group categorizations) supporting the robustness of our primary findings (eTable 2 and eTable 3).

This study has several limitations. First, we acknowledge that this cross-sectional design precludes inference about temporal relationships or causality. Second, our study focused on global structural brain volumes rather than focal, microstructural, or functional brain markers and subsequently may not fully capture subtler or region-specific neurobiological differences between groups. Third, we used a conservative multivariable models that excluded our study participants (mean age of 58.5 years old) with baseline dementia, cerebrovascular disease, and cardiovascular disease with additionally adjusted for baseline ARWMC (to account for cerebral small vessel disease) within mediation framework. These choices may yield more conservative estimates of associations and indirect effects through global brain volumes, and thus the non-significant associations should be interpreted cautiously.

In conclusion, higher CAIDE score was independently associated with poorer cognitive performance in this middle-to-older aged population. Overall structural brain volumes (including GM, WM and CSF volume) explained only a small proportion of these associations, suggesting that macroscopic brain atrophy alone may not fully explain the observed cognitive vulnerability associated with elevated CAIDE score. These findings suggest that additional biological processes beyond global structural brain measures may contribute to cognitive vulnerability associated with CAIDE score, potentially reflecting broader biological vulnerability.

## Supporting information

eAppendix

## Author contributions

our corresponding authors had full access to all of the data in the study and took responsibility for the integrity of data and the accuracy of the data analysis. Concept and design G. Shin, A Siddiquee, June Kang, N. Kim, C. Shin. Acquisition, analysis, or interpretation data G. Shin, A Siddiquee, S. Lee, B. Kim, and Y.Kim. Drafting the manuscript: G. Shin. Critical revision of manuscript for important intellectual content All authors. Statistical Analysis: G. Shin. Obtained funding, N. Kim., Administrative, technical, or material support, G. Shin, June Kang, S. Lee, C. Shin, and N. Kim.

## Role of the funder or sponsor

The funders had no role in the design and conduct of the study; collection, management, analysis, and interpretation of the data; preparation, review, or approval of the manuscript; and decision to submit the manuscript for publication.

## Data Availability

This data is not for public and can only be used (de-identified participant data) by those who included in IRB plan. A contact can be made by following email address

https://nih.go.kr/ko/main/contents.do?menuNo=300566

## Acknowledgements

This study was performed with bio-resources from National Biobank of Korea, the National Institute of Health (NIH), Republic of Korea. Genotype data were provided by the Collaborative Genome Program for Fostering New Post-Genome Industry (3000-3031b). We sincerely thank Dr. Daniela Enache for helpful comments and valuable feedback on our manuscript.

