## Supplementary material for "CAIDE score, brain structure, and cognitive functions in middle-to-older aged adults: A KoGES population-based study": eAppendix

**Supplement Appendix**

Shin GS, Siddiquee AT, et al. CAIDE risk, brain structure, and cognition in middle-to-late life: a KoGES population Study.

eFgiure 1. Pearson correlation matrix of CAIDE components, structural brain volumes, and neuropsychological assessment battery (n=2,425).

eTable 1. (a) CAIDE score components without (N=2,425) or with (N=2,063) APOE4 carrier status in middle-to-older aged population and (b) distribution of CAIDE score study participants without and with APOE4 carrier status.

eTable 2. Multivariable adjusted β estimates with 95% Cl for associations between CAIDE risk levels (a total up to 18 points by including APOE4 carrier status) and cognitive performance (N = 2,063).

eTable 3. Multivariable adjusted β estimates for associations between CAIDE risk levels (low and moderate risk vs. high risk) with 95% Cl (N = 2,425).

eFigure 1. Pearson correlation matrix of CAIDE components, structural brain volumes, and neuropsychological assessment battery (n=2,425).

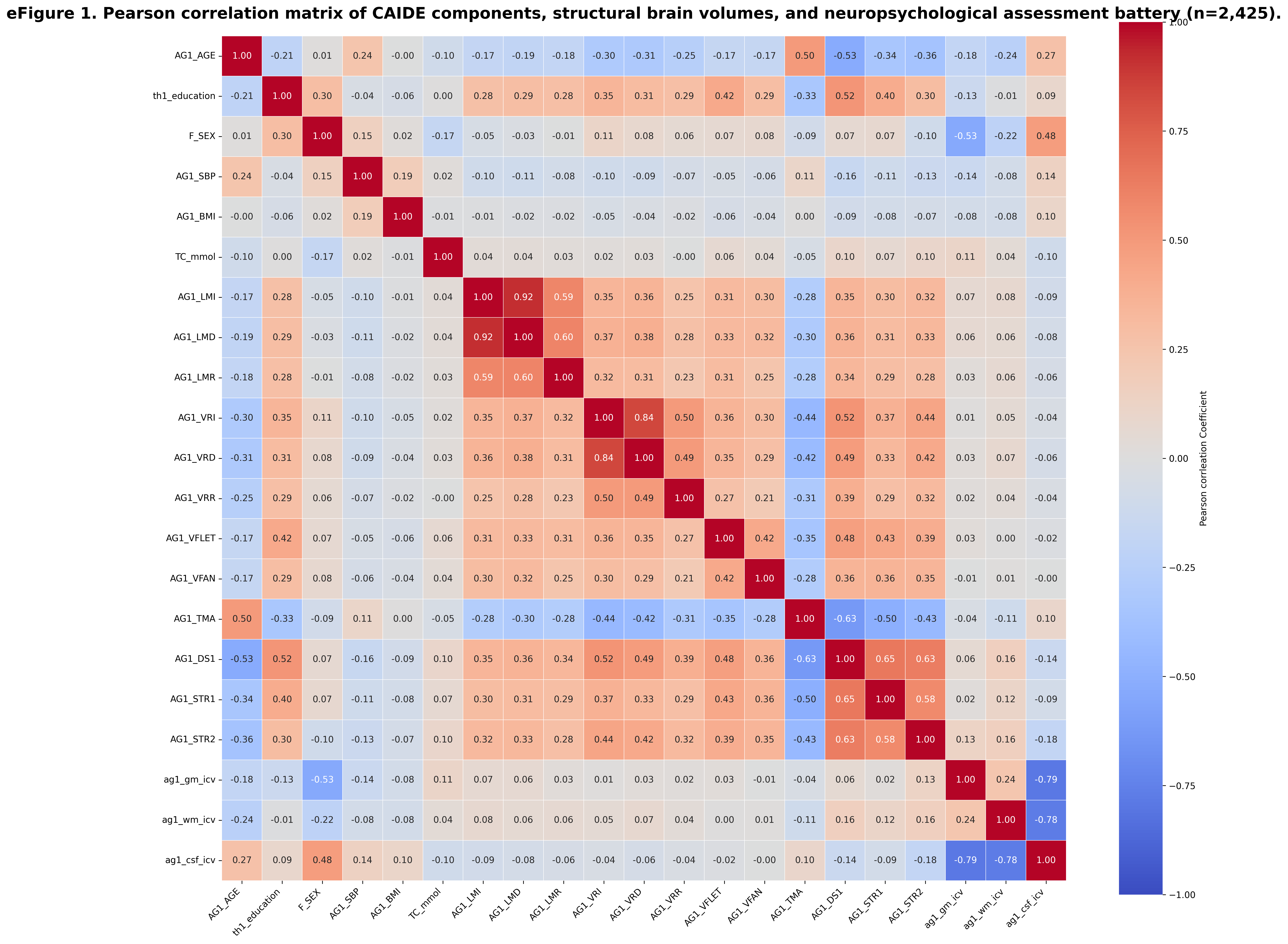

eTable 1a. CAIDE score components with/without APOE4 carrier status in middle-to-older aged population.

| CAIDE factors | N=2,425^†^ | | N=2,063^††^ | |
| --- | --- | --- | --- | --- |
|  | Measurements | Points | Measurements | Points |
| Age (years) | age < 47 yrs | 0 | age < 47 yrs | 0 |
|  | 47 yrs ≤ age ≤ 53 yrs | 3 | 47 yrs ≤ age ≤ 53 yrs | 3 |
|  | 53 yrs < age | 4 | 53 yrs < age | 5 |
| Sex (M/F) | men | 1 | men | 1 |
|  | women | 0 | women | 0 |
| Education (levels) | middle school < | 0 | middle school < | 0 |
|  | middle school | 2 | middle school | 3 |
|  | < middle school | 3 | < middle school | 4 |
| ^‡^Body Mass Index (kg/m^2^) | BMI ≥ 25 | 2 | BMI ≥ 25 | 2 |
|  | BMI < 25 | 0 | BMI < 25 | 0 |
| Physical Activity | < 2 times/week, | 1 | < 2 times/week, | 1 |
|  | ≥30 min/session |  | ≥30 min/session |  |
| (times/week, minutes/session) | ≥2 times/week, | 0 | ≥2 times/week, | 0 |
|  | ≥30 min/session |  | ≥30 min/session |  |
| Total Cholesterol (mmol/L) | TC > 6.5 mmol/L | 2 | TC > 6.5 mmol/L | 1 |
|  | 6.5 mmol/L ≥ TC | 0 | 6.5 mmol/L ≥ TC | 0 |
| Systolic Blood Pressure (mmHg) | SBP > 140 mmHg | 2 | SBP > 140 mmHg | 2 |
|  | SBP ≤ 140 mmHg | 0 | SBP ≤ 140 mmHg | 0 |
| Alipoprotein E4 carrier | carrier | N/A | carrier | 2 |
|  | no carrier | N/A | no carrier | 0 |
| Total score | 15 | | 18 | |

‡ Korean cut-off for obesity (KSSO)

† Overall study participants regardless of APOE4 genotype data †† included eligible APOE genotype data

eTable 1b. Distribution of CAIDE score study participants with/without APOE4 carrier status

| **APOE4 status** | **CAIDE score** | **number of participants (n)** | **percentages (%)** |
| --- | --- | --- | --- |
| without APOE4^†^ | low (0-5) | 808 | 33.3 |
|  | moderate (6-7) | 945 | 38.9 |
|  | high (7<) | 672 | 27.7 |
| with APOE4^††^ | low (0-6) | 663 | 32.1 |
|  | moderate (7-8) | 647 | 31.6 |
|  | high (8<) | 753 | 36.5 |

† Overall study participants regardless of APOE4 genotype data †† included eligible APOE genotype data

eTable 2. Multivariable adjusted β estimates with 95% Cl for associations between CAIDE risk levels (a total up to 18 points by including APOE4 carrier status) and cognitive performance (N = 2,063).

|  |  | CRUDE | | Adjusted Model | |
| --- | --- | --- | --- | --- | --- |
|  |  | Estimate (95% CI) | p-value | Estimate (95% CI) | ^†^p-value |
| Story recall tests |  |  |  |  |  |
| Moderate Risk | Immediate recall | -0.145 (-0.272 to -0.017) | 0.021 | -0.141 (-0.269 to -0.012) | 0.028 |
|  | Delayed recall | -0.134 (-0.261 to -0.007) | 0.036 | -0.128 (-0.256 to 0.001) | 0.051 |
|  | Recognition | -0.152 (-0.279 to -0.025) | 0.014 | -0.137 (-0.265 to -0.009) | 0.033 |
| High Risk | Immediate recall | -0.423 (-0.546 to -0.301) | <.001 | -0.405 (-0.530 to -0.280) | <.001 |
|  | Delayed recall | -0.469 (-0.591 to -0.346) | <.001 | -0.441 (-0.565 to -0.316) | <.001 |
|  | Recognition | -0.424 (-0.547 to -0.301) | <.001 | -0.382 (-0.506 to -0.257) | <.001 |
| Visual Reproduction tests |  |  |  |  |  |
| Moderate Risk | Immediate recall | -0.066 (-0.192 to 0.059) | 0.437 | -0.076 (-0.201 to 0.547) | 0.333 |
|  | Delayed recall | -0.073 (-0.199 to 0.052) | 0.351 | -0.079 (-0.205 to 0.046) | 0.296 |
|  | Recognition | -0.097 (-0.224 to 0.030) | 0.174 | -0.097 (-0.224 to 0.031) | 0.176 |
| High Risk | Immediate recall | -0.532 (-0.654 to -0.411) | <.001 | -0.501 (-0.623 to -0.379) | <.001 |
|  | Delayed recall | -0.551 (-0.672 to -0.429) | <.001 | -0.512 (-0.634 to -0.391) | <.001 |
|  | Recognition | -0.467 (-0.589 to -0.345) | <.001 | -0.438 (-0.562 to -0.314) | <.001 |
| Verbal Fluency Tests |  |  |  |  |  |
| Moderate Risk | Phonemic | -0.082 (-0.208 to 0.043) | 0.275 | -0.090 (-0.216 to 0.036) | 0.212 |
|  | Category | -0.042 (-0.170 to 0.086) | 0.724 | -0.044 (-0.171 to 0.084) | 0.702 |
| High Risk | Phonemic | -0.526 (-0.647 to -0.404) | <.001 | 0.495 (-0.617 to -0.373) | <.001 |
|  | Category | -0.370 (-0.493 to -0.247) | <.001 | -0.325 (-0.449 to -0.201) | <.001 |
| Trail making Test |  |  |  |  |  |
| Moderate Risk | Trails A time | 0.156 (0.032 to 0.281) | 0.009 | 0.155 (0.032 to 0.278) | 0.009 |
| High Risk | Trails A time | 0.629 (0.509 to 0.749) | <.001 | 0.569 (0.450 to 0.689) | <.001 |
| Digit symbol test |  |  |  |  |  |
| Moderate Risk | Coding | -0.237 (-0.356 to -0.119) | <.001 | -0.227 (-0.344 to -0.110) | <.001 |
| High Risk | Coding | -0.927 (-1.041 to -0.812) | <.001 | -0.859 (-0.973 to -0.745) | <.001 |
| Stroop tests |  |  |  |  |  |
| Moderate Risk | Word reading | -0.188 (-0.312 to -0.065) | 0.001 | -0.179 (-0.302 to -0.056) | 0.002 |
|  | Color reading | -0.239 (-0.364 to -0.113) | <.001 | -0.187 (-0.311 to -0.063) | 0.001 |
| High Risk | Word reading | -0.716 (-0.834 to -0.597) | <.001 | -0.663 (-0.782 to -0.543) | <.001 |
|  | Color reading | -0.628 (-0.749 to -0.508) | <.001 | -0.542 (-0.663 to -0.422) | <.001 |

*Unadjusted

‡Fully Adjusted: adjusted for baseline smoking, drinking, BDI, DM, and ARWMC.

† post-hoc Tukey

eTable 3. Multivariable adjusted β estimates for associations between CAIDE risk levels (low and moderate risk vs. high risk) with 95% Cl (N = 2,425).

|  |  | CRUDE | | Adjusted Model | |
| --- | --- | --- | --- | --- | --- |
|  |  | Estimate (95% CI) | p-value | Estimate (95% CI) | ^†^p-value |
| Story recall tests |  |  |  |  |  |
| High Risk | Immediate recall | -0.366 (-0.453 to -0.278) | <.001 | -0.351 (-0.439 to -0.263) | <.001 |
|  | Delayed recall | -0.396 (-0.483 to -0.308) | <.001 | -0.376 (-0.464 to -0.288) | <.001 |
|  | Recognition | -0.355 (-0.443 to -0.267) | <.001 | -0.329 (-0.418 to -0.242) | <.001 |
| Visual Reproduction tests |  |  |  |  |  |
| High Risk | Immediate recall | -0.496 (-0.582 to -0.409) | <.001 | -0.479 (-0.565 to -0.393) | <.001 |
|  | Delayed recall | -0.467 (-0.554 to -0.380) | <.001 | -0.445 (-0.531 to -0.359) | <.001 |
|  | Recognition | -0.460 (-0.547 to -0.373) | <.001 | -0.444 (-0.531 to -0.357) | <.001 |
| Verbal Fluency Tests |  |  |  |  |  |
| High Risk | Phonemic | -0.473 (-0.560 to -0.386) | <.001 | -0.452 (-0.539 to -0.366) | <.001 |
|  | Category | -0.352 (-0.439 to -0.264) | <.001 | -0.325 (-0.412 to -0.238) | <.001 |
| Trail making Test |  |  |  |  |  |
| High Risk | Trails A time | 0.513 (0.426 to 0.599) | <.001 | 0.477 (0.392 to 0.562) | <.001 |
| Digit symbol test |  |  |  |  |  |
| High Risk | Coding | -0.774 (-0.857 to -0.690) | <.001 | -0.736 (-0.818 to -0.654) | <.001 |
| Stroop tests |  |  |  |  |  |
| High Risk | Word reading | -0.615 (-0.700 to -0.529) | <.001 | -0.583 (-0.668 to -0.498) | <.001 |
|  | Color reading | -0.474 (-0.561 to -0.387) | <.001 | -0.428 (-0.513 to -0.342) | <.001 |

*Unadjusted

‡Fully Adjusted: adjusted for baseline smoking, drinking, BDI, DM, and ARWMC.

† post-hoc Tukey
